# Trauma Exposure, PTSD, and Methylation of the Blood Brain Barrier Claudin-5 Gene

**DOI:** 10.1101/2025.11.24.25340875

**Authors:** Erika J. Wolf, Xiang Zhao, Annelise Madison, Jack Carbaugh, Catherine B. Fortier, William P. Milberg, Traumatic Stress Brain Research Group, Mark W. Logue, Mark W. Miller

## Abstract

Posttraumatic stress disorder (PTSD) is associated with early onset of neurological conditions, but the mechanism by which PTSD relates to diseases of the central nervous system is unclear. One possibility is that PTSD perpetuates breakdown of the blood brain barrier (BBB), allowing for bidirectional passage of molecules across the periphery and central nervous system that promote neuropathology. Preclinical studies have implicated claudin-5 (CLDN5), a protein integral to the integrity of the BBB tight junctions, in the pathogenesis of depression. Based on this, we evaluated if trauma exposure and PTSD related to *CLDN5* epigenetics in blood among 1,311 trauma-exposed individuals (primarily Veterans) and in the brain tissue from 100 decedents. Three (out of 19) *CLDN5* DNA methylation (DNAm) probes, cg00804504, cg17411190, and cg21872764, were significantly associated with trauma exposure or PTSD severity after multiple testing correction in blood. The latter two probes also showed association with PTSD diagnosis in ventromedial prefrontal cortex. The most strongly associated DNAm probe, cg21872764, also evidenced associations with the neuropathology biomarker neurofilament light in plasma. *CLDN5* expression was strongly associated with estimated proportion of brain endothelial cells. The cross-sectional associations observed in this study highlight the importance of studying the link between traumatic stress and early onset of neuropathology. Future research is needed to test the mechanistic hypothesis that trauma exposure and chronic PTSD alter *CLDN5* DNAm, lead to increased BBB permeability and allow for bidirectional passage of neuroinflammatory molecules across the BBB.

## 1. Introduction

Posttraumatic stress disorder (PTSD) is associated with risk for early onset of neurodegeneration (Miller & Sadeh, 2014), declining cognitive function (Roberts et al., 2022), and dementia (Yaffe et al., 2010). Among older Veterans who receive VA healthcare, PTSD is associated with approximately double the risk for a dementia diagnosis compared to individuals with no lifetime history of PTSD (Yaffe et al. 2010, Logue et al., 2022). These neurological alterations among individuals with PTSD may partially reflect an accelerated cellular aging process (Sugden et al., 2022) whereby the stress of chronic psychiatric symptoms, such as emotional and physiological reactivity, sleep disturbance, and associated poor health behaviors, lay the groundwork for advanced biological aging (Miller, et al., 2018; Bourassa & Sbarra, 2024). It is unclear how psychiatric symptoms impact central nervous system (CNS) structures and processes or how psychopathology-related aberrations in peripheral biomarkers (e.g., inflammation, glucocorticoid reactivity) relate to alterations in the CNS (e.g., neuroinflammation, neurodegeneration) and vice versa. One possibility is that psychiatric stress and associated physiological reactivity perpetuate breakdown of the blood brain barrier (BBB), allowing for bidirectional passage of molecules that promote neuroinflammation and neuropathology (Medina-Rodriguez & Beurel, 2022; Wu et al., 2022; Kealy et al., 2020).

### 1.1 Claudin-5, the Blood Brain Barrier, and Neurological Disease

The BBB serves to limit molecules from passing between the CNS and the periphery so that only select molecules can cross, which protects the CNS and maintains homeostasis. This is accomplished through a layer of tightly connected endothelial cells aligned along the capillaries of the brain that separate blood vessels from the CNS. The BBB is part of a larger structure referred to as the neurovascular unit, comprised of astrocytes, microglia, mural and endothelial cells, and neurons, which coordinates cellular signaling and allows blood, nutrients, neuroimmune molecules, and CNS waste to cross the BBB (McConnell & Mishra, 2022). The endothelial cells of the neurovascular unit are connected to each other through tight junction proteins which form a physical barrier (akin to a seal) and regulate passive transfer of molecules across it. Several types of claudin and occludin proteins control the membranes at the tight junctions. Chief among these is claudin-5 (CLDN5), which is expressed primarily in brain endothelial cells and is critical to tight junction integrity and resistance to small molecule transfer across the BBB (Greene et al., 2019; Hashimoto et al., 2023).

Numerous factors alter *CLDN5* expression, including inflammatory proteins such as tumor necrosis factor alpha (TNFα) and associated nuclear factor kappa B signaling, glucocorticoids, and circadian-related proteins (Greene et al., 2019). Downregulation of *CLDN5* is associated with increased BBB leakiness and passage of peripheral immune and inflammatory cells into the CNS, which can be neurotoxic (Greene et al., 2019). *CLDN5* downregulation is also associated with movement of central neuropathology biomarkers into the periphery. For example, downregulation of *CLDN5* in the brain allows for transfer of amyloid beta (Aβ), a primary indicator of Alzheimer’s disease (Jack et al., 2024), from the CNS into the periphery via the BBB; Aβ itself may autoregulate *CLDN5* expression to allow for its clearance from the brain (Keaney et al., 2015). Neurofilament light (NFL), a cytoskeletal protein produced in neuronal cytoplasm, is a marker of axonal injury that is correlated with neuropathological disease (Jack et al., 2024; Yuan & Nixon, 2021). It can be measured in the CNS and is also detectable in plasma and serum (from cerebral spinal fluid; Yuan & Nixon, 2021). NFL is also associated with BBB permeability (Friis et al., 2022) and CLDN5 levels (Li et al., 2021), so its presence in the periphery may be a sign of increased BBB permeability. Another neurology protein with relevance to CLDN5 is glial fibrillary acidic protein (GFAP), which forms part of the astrocyte cytoskeleton structure. GFAP-immunoreactive astrocytes are reduced in concert with alterations in CLDN5 protein levels and organization (Camire et al., 2015; Sántha et al., 2015) and GFAP levels are relevant to dementia risk (Kim et al., 2023; Gonzales et al., 2022). Alterations in Aβs, NFL, and GFAP may reflect, in part, the underlying role of *CLDN5* in BBB disruption and neurological disease. Consistent with this, reduced *CLDN5* expression and associated disruption to the BBB at the tight junctions is associated with numerous neurological conditions including traumatic brain injury sequela, Alzheimer’s disease, cognitive decline, multiple sclerosis, epilepsy, and stroke (Greene et al., 2019; Hashimoto et al., 2023).

### 1.2 Claudin-5 and Stress-Related Psychopathology

In addition to associations with neurological conditions, *CLDN5* is also relevant to stress-related psychopathology. Much of the evidence for this comes from rodent models of depression. Chronic social stress is associated with reduced cldn5 expression in the nucleus accumbens of stress-susceptible male mice and in the prefrontal cortex of stress-susceptible female mice, and this co-occurs with depression-like phenotypes and increased BBB permeability (Menard et al., 2017; Dion-Albert et al., 2022). Both the reduced cldn5 expression in the nucleus accumbens and the depression-like phenotype can be reversed with lengthy administration of the antidepressant imipramine (Menard et al., 2017). Rodent studies also suggest that acute and chronic stress are related to reduced cldn5 expression in the hippocampus, loss of regional BBB integrity, and behavioral and cognitive impairments (Menard et al., 2017; Sun et al., 2024; Ni et al., 2022). Experimentally manipulated hippocampal cldn5 downregulation was also associated with increased depression-like behaviors in rodents and reversed with long term administration of fluoxetine (Sun et al., 2024). Stress-related cldn5 downregulation may allow for inflammatory molecules to pass into the CNS: experimentally stressed mice with selectively downregulated cldn5 expression showed depression-like phenotypes and infiltration of the inflammatory molecules interleukin (IL)-6 and TNFα from the periphery into the nucleus accumbens (Menard et al., 2017) and hippocampus (Sun et al., 2024), respectively. Secretion of TNFα and IL-6 by microglia may also be sufficient to damage the BBB via reduced expression of *CLDN5* per *in vitro* studies (Camire et al., 2015). Together, this raises the possibility of both central and peripheral inflammation leading to tight junction damage.

Postmortem human studies also suggest that *CLDN5* expression is substantially reduced in the nucleus accumbens from untreated depressed men and women (Menard et al., 2017), and in the ventromedial prefrontal cortex (vmPFC) of women with depression (Dion-Albert et al., 2022). Similarly, postmortem studies suggest that CLDN5 protein is reduced in the hippocampi of men and women with depression and schizophrenia, with greater reductions in *CLDN5* expression as a function of more years of chronic psychiatric symptoms (Greene et al., 2020).

### 1.3 *CLDN5* Genotypes and DNA Methylation

A handful of studies have examined *CLDN5* candidate genotypes in association with psychiatric outcomes. The minor allele of the rs10314 *CLDN5* variant may increase risk for schizophrenia (Greene et al., 2018; Sun et al., 2004). A UK Biobank candidate SNP study with over 275k individuals found that *CLDN5* variant rs885985 interacted with an IL-6 variant and self-reported life stress to predict depression (i.e., a 3-way interaction effect), and this was replicated in a second cohort (Gal et al., 2023). Associations between these genotypes and PTSD risk have not been evaluated in candidate gene studies, though the most recent genome wide association study of PTSD featuring over 1.2 million participants did not report a significant association with either variant (*p*s ≥ .20; Nievergelt et al., 2024).

Given the evidence for *CLDN5* genotypes and expression in BBB integrity and stress-associated psycho- and neuropathology, it is reasonable to wonder about the role of DNA methylation (DNAm) in this process, particularly among aging populations. This is because DNAm is responsive to the environment and physiological processes (Tobi et al., 2009), known to change with age (Christensen et al., 2009), and is a primary driver of gene expression. It may serve as the environmentally sensitive intermediate process by which substantial stressors like chronic PTSD symptoms exert effects on BBB.

To our knowledge, the only *CLDN5* DNAm study relevant to understanding neuropathology stems from a longitudinal cognitive trajectory and brain donation study with a cohort of over 600 individuals (Hüls et al., 2022). That study found that cognitive trajectory was strongly (at the genome-wide significance level) associated with postmortem DNAm at two *CLDN5* loci in the dorsolateral prefrontal cortex (dlPFC). The probes, cs16773741 and cg05460329, were associated with greater cognitive decline over time among individuals with and without obvious neuropathology. Though the results supported a role for *CLDN5* DNAm in cognitive outcomes, associations between DNAm and psychopathology were not examined.

### 1.4 Study Aims & Hypotheses

Given evidence for a role for *CLDN5* in susceptibility to depression following stress (Menard et al., 2017), and phenotypic and genetic similarities between PTSD and depression (Dalvie et al., 2021; Ringwald et al., 2023), our aim was to examine, for the first time, the association between PTSD and *CLDN5* epigenetics. Our primary hypothesis was that chronic PTSD symptoms would be associated with alterations in *CLDN5* DNAm. We first examined this in blood DNAm in a cohort of over 1300 trauma-exposed individuals who were assessed for PTSD using structured diagnostic interviews. We also evaluated if *CLDN5* candidate genotypes accounted for associations between PTSD and *CLDN5* DNAm. We then tested our primary hypothesis in postmortem brain tissue samples from 100 donors by examining associations between PTSD diagnosis and *CLDN5* DNAm in vmPFC, dlPFC, and motor cortex. The postmortem tissue afforded the opportunity to examine associations between *CLDN5* DNAm and expression and between *CLDN5* expression and estimated brain endothelial cell proportions.

Our second hypothesis was that *CLDN5* DNAm would be associated with peripheral measures of neuropathology and inflammation. We examined cross-sectional associations between *CLDN5* DNAm and the neuropathology biomarkers Aβ-42/Aβ-40 (i.e., their ratio), NFL, GFAP, and the inflammatory biomarkers IL-6 and TNFα, all measured in blood in our trauma-exposed cohort. As reviewed above, these were selected based on their relevance to CLDN5 (Keaney et al., 2015), aging and neurodegeneration (Jack et al., 2024), and PTSD (Wolf et al., 2024; Miller et al., 2024; Peruzzolo et al., 2022). Evidence for these associations could be a reflection of a leaky BBB allowing for neuropathology biomarkers to pass from the brain to blood.

## 2. Methods

### 2.1 Participants and Procedures

Participants (*N* = 1311) were drawn from two cohorts and the data were combined for analyses. The National Center for PTSD (NCPTSD) cohort (*n* = 752) included trauma-exposed male (60.6%) and female Veterans from a range of war eras (i.e., WWII through post-9/11) and a subset of their live-in partners with trauma exposure (*n* = 186) who underwent identical procedures (Wolf et al., 2024, Logue et al., 2013). On average, the NCPTSD cohort was in their early 50s (range: 19 to 75 years). The Translational Research Center for TBI and Stress Disorders (TRACTS) cohort (*n* = 559) included male (90.3%) and female post-9/11 combat-exposed Veterans with a mean age in their early 30s (McGlinchey et al., 2017). Both cohorts included overlapping assessment and blood draw procedures. The demographic characteristics of the combined cohort are listed in Table 1.

**Table 1.** Demographic Characteristics of the Cohorts.

| Variable | Human Subjects (*N* = 1311) |  | Brain Bank (*N* = 100) |  |
| --- | --- | --- | --- | --- |
|  | % (*n*) | *M* (*SD*) | % (*n*) | *M* (*SD*) |
| Sex (male) | 76.1 (998) |  | 60.0 (60) |  |
| Age |  | 44.24 (13.68) |  | 43.70 (12.93) |
| Veteran | 85.8 (1125) |  | 22.0 (22) |  |
| Ethnicity: Hispanic or Latino/a | 16.2 (213) |  | NA |  |
| Race |  |  |  |  |
| White | 76.8 (1007) |  | 76.0 (76) |  |
| Black | 13.0 (171) |  | 23.0 (23) |  |
| American Indian | 5.0 (66) |  | NA |  |
| Asian | 1.8 (23) |  | NA |  |
| Other | 8.6 (113) |  | 1.0 (1) |  |
| Current PTSD Diagnosis | 48.2 (632) |  | 40.0 (40) |  |
| Current PTSD Severity |  | .33 (.22) |  | NA |
| Lifetime Trauma Count |  | 3.14 (2.40) |  | NA |

*Note.* Ethnicity and race were self-reported and options were not mutually exclusive; percentages were combined into “other” for reporting purposes due to small cell sizes for some response options such as Hawaiian/Pacific Islander, other, and unknown. PTSD severity reflects the harmonized PTSD severity score that captures symptom severity ratings a percentage of the maximum possible severity score. Some characteristics were not available in the brain bank cohort and are labeled as NA (not applicable).

Across the two cohorts, participants were recruited from VA PTSD clinics, research recruitment databases and flyers, and military and Veteran community events. Exclusion criteria included acute substance intoxication and acute psychotic or safety (suicidal and homicidal) concerns. Additionally, the TRACTS protocol excluded Veterans with a neurological or cognitive disorder diagnosis other than those related to traumatic brain injury (TBI). Participants in both cohorts completed psychological interviews, self-report surveys, blood draw, and physiological measurements. The studies were approved by the VA Boston Healthcare System IRB and all participants provided written informed consent.

### 2.2 Measures

In the NCPTSD cohort, current PTSD was assessed with the Clinician Administered PTSD Scale for *DSM-IV* (CAPS; Blake et al., 1995) or *DSM-5* (CAPS-5; Weathers et al., 2018), based on the prevailing version of the *DSM* at the time of study participation. These interviews were videotaped and approximately 25% were reviewed by an independent rater. Intraclass correlation coefficients for PTSD symptom severity totals across two timepoints were *r* ≥ .84 and diagnostic agreement for current PTSD diagnoses were Κ ≥ .87. For the TRACTS cohort, the CAPS for *DSM-IV* was administered and all interviews were audio recorded; 23 were reviewed by an independent rater which yielded intraclass correlation *r* = .92 for PTSD symptom severity and Κ = .68 for PTSD diagnostic status. To harmonize current PTSD severity scores (sum of all CAPS PTSD items) across the two versions of the measure (which have different possible ranges), we calculated a standardized score which reflected the total symptom severity score as a fraction of the maximum possible total score on each measure. These scores have a potential range of 0 to 1. Trauma exposure was assessed with the self-report Traumatic Life Events Questionnaire (TLEQ; Kubany et al., 2000) which assesses exposure to 23 types of traumatic experiences. We tabulated the number of different types of traumatic events endorsed for use in our analyses. Comorbid major depressive disorder (MDD) diagnoses were assessed with the Structured Clinical Interview for *DSM-IV* (First et al., 1994) or *DSM-5* (First et al., 2015) Disorders.

### 2.3 Biomarkers

#### 2.3.1 DNA and DNAm

Fasting morning peripheral blood samples were drawn into 10 mL EDTA tubes which were centrifuged the same day. Plasma, serum, and buffy coat samples were stored at -80° C until ready for analysis. DNA was extracted from buffy coat for genotype and DNAm ascertainment using Qiagen reagents on a Qiagen Gentra AutoPure machine. Genotypes were interrogated on the Illumina HumanOmni2.5-8 Beadchip. DNAm was obtained on the Illumina Infinium EPIC Beadchip. DNA and DNAm plates were balanced for sex and PTSD diagnostic status. Details of the genotyping data quality control (QC) and imputation procedures can be found in the Supplementary Materials. We computed ancestry (for use in the DNAm analyses) and ancestry substructure (within the EUR ancestry subset, for use in the genotype analyses) principal components (PCs) from 100,000 randomly chosen genotypes with minor allele frequency (MAF) of ≥ 5% (Nievergelt et al., 2019).

We followed a pipeline developed by the PGC PTSD Epigenetics Working Group (Ratanatharathorn et al., 2017) and its updated version at https://github.com/PGC-PTSD-EWAS/EPIC_QC to process DNAm data (Supplementary Materials). Proportional estimates of six DNAm-based cell types (B cells, CD4+T cells, CD8+T cells, natural killer cells, monocytes, and neutrophils) were computed using the IDOL algorithm (Koestler et al., 2016) in the Bioconductor package EpiDISH (Teschendorff et al., 2017) in R. A DNAm-based smoking score was calculated by taking the product of the methylation M values at 39 smoking-associated CpG sites and the effect size estimates of their association with smoking pack years (Li et al., 2018).

#### 2.3.2 Simoa Neuropathology & Inflammatory Biomarkers

The following Simoa analytes were analyzed from peripheral plasma samples in the NCPTSD and TRACTS cohorts: IL-6, TNFα, GFAP, NFL, Aβ-42, and Aβ-40 (the latter two were combined into a single variable reflecting the ratio of Aβ-42 to Aβ-40; Lewczuk et al., 2004). As previously described (Wolf et al., 2024; Miller et al., 2024), Simoa assays for the NCPTSD cohort were obtained directly from the Quanterix Accelerator Lab (Quanterix Corporation, Billerica, MA; see Supplementary Materials for QC and other details). In the TRACTS cohort, the plasma samples were analyzed (in duplicate) on an in-house HD-1 analyzer (Quanterix, Billerica, MA; see Supplementary Materials).

### 2.4 Postmortem Brain Bank Cohort

Data from the VA National PTSD Brain Bank cohort were from *n* = 117 donors originally collected by the Lieber Institute for Brain Development at Johns Hopkins University (Friedman et al., 2017; Mighdoll et al., 2018). Left hemisphere postmortem brains were acquired and three brain regions relevant to PTSD and aging were available to our group: dlPFC, vmPFC, and motor cortex. We have previously described the methods for obtaining these regions and ascertaining genotypes, DNAm, and RNA sequence data (Wolf et al., 2021; Zhao et al., 2022). This cohort excluded individuals with neurodegenerative disease, neuritic pathology, or severe TBI as determined by board-certified neuropathologists. Determination of psychiatric diagnoses was based on medical record review and next-of-kin interviews performed by board-certified neuropathologists. The interviews included the PTSD Checklist for DSM-5 adapted for postmortem use, the MINI International Neuropsychiatric Interview 6.0, and the Liber Psychological Autopsy Interview (Mighdoll et al., 2018). Two independent board-certified psychiatrists reviewed the records and psychiatric diagnostic determinations and made confidence ratings for the assigned diagnoses using a 1-5 scale. PTSD cases had confidence ratings of 3 or greater.

DNAm was obtained on the Illumina Infinium MethylationEPIC beadchip with processing and QC procedures as described for plasma cells and in previous publications (Logue et al., 2020). The proportion of neurons was estimated from the methylation data using the CETS package (Guintivano et al., 2013). RNA was obtained from 25 mg of tissue via Qiagen RNeasy Fibrous Tissue Minikit. Illumina TruSeq Stranded total RNA kit with globin depletion and a Hiseq 2500 for paired-end 75bp reads for library sequencing. RNA integrity values were available for a subset of samples and found to be acceptable. Quality surrogate variables (qSVs) were generated to assess RNA degradation using the Bioconductor package sva (Leek et al., 2012). Samples were excluded if < 50% uniquely mapped reads or if they were outliers per evaluation of log-transformed counts from the regularized log transformations in DESeq2 (Love et al., 2014). Estimated proportional cell types (astrocytes, endothelial cells, microglia, mural cells, neurons, oligodendrocytes, and red blood cells) were generated for each brain region per BrainInABlender (Hagenauer et al., 1995).

We used the largest possible dataset for each set of brain bank analyses. For analyses examining associations between PTSD and *CLDN5* DNAm across brain regions, decedents were excluded if they had a bipolar disorder diagnosis (*n* = 5). Among the remaining 112 donors, *n* = 42 had PTSD (*n* = 35 of this group had comorbid MDD), *n* = 41 had MDD without PTSD and *n* = 29 were controls. Given our efforts to replicate associations with PTSD from our living cohort, we excluded the cases with MDD without PTSD. We further excluded two individuals missing antidepressant use data and one motor cortex sample that failed DNAm QC, yielding a final sample size of *n* = 69 with dlPFC and vmPFC data, and *n* = 68 for motor cortex data for the DNAm and PTSD analyses.

For the expression analyses, we focused on associations between biological variables (e.g., *CLDN5* expression with estimated cell types, *CLDN5* DNAm with expression) and did not include psychological diagnoses in the analyses. Therefore, we included all subjects regardless of MDD status (bipolar cases were still excluded). RNA was available for a subset of the non-bipolar donors (*n* = 92-96 across brain regions). Individuals whose RNA data failed QC procedures (*n* = 2 - 5 across brain regions) or had missing genotypes for determining ancestry (*n* = 1) were removed from expression analyses. This yielded a sample size of *n* = 93 for dlPFC, 86 for vmPFC, and 89 for motor cortex. Collectively, data from 100 donors were included in either DNAm and PTSD or expression-related brain bank analyses, and their demographic characteristics are listed in Table 1.

### 2.5 Data Analyses

All analyses involved multiple linear regression models conducted in SPSS v 29.0.1.0 (IBM Corporation) or R version 4.0.5. We first evaluated the 19 *CLDN5* DNAm probes (the outcome variable) on the EPIC chip that surpassed QC in association with current PTSD symptom severity in our largest possible cohort (*N* = 1311). These regressions covaried for age, sex, five WBCs (CD8 and CD4-T cells, monocytes, b cells, and natural killer cells), and the top 3 ancestry PCs. We employed a false discovery rate (FDR; Benjamini & Hochberg, 1995) adjusted *p*-value (*p*_-adj_) threshold of *p*_-adj_ < .05. Sensitivity analyses which further covaried for smoking (using the DNAm smoking score), lifetime trauma exposure, and MDD were conducted for models with significant PTSD associations with a given probe. Next, we examined the candidate *CLDN5* variants rs10314 and rs885985, for their associations with the PTSD-associated *CLDN5* loci to determine if they accounted for associations between PTSD and *CLDN5* DNAm (in regressions covarying for age, sex, WBCs, and 3 ancestry substructure PCs). This was conducted in the EUR subset (*n* = 873) due to concerns of population stratification. We also examined the SNP associations with trauma exposure and PTSD severity in analogous regression models. To test our hypothesis that *CLDN5* DNAm alterations would relate to increased levels of neurology and inflammation biomarkers in blood, we examined *CLDN5* DNAm loci (that were significantly related to PTSD) in association with peripheral IL-6, TNFα, GFAP, Aβ-42/Aβ-40, and NFL while covarying for sex, age, and WBCs and employing FDR *p*-value adjustments for the five Simoa biomarkers (*n* = 1097 of mixed ancestry due to missing Simoa data).

Finally, we examined data derived from postmortem brain tissue to test for replication of results. In the three brain regions, we tested associations between PTSD diagnosis (versus controls) and the *CLDN5* loci that were identified as being significantly associated with PTSD in blood, covarying for age at death, sex, postmortem interval (PMI), 3 PCs, proportion of neurons, and antidepressant use (Menard et al., 2017). For completeness, we also tested for association between PTSD and all 19 *CLDN5* DNAm probes on the chip in each brain region. We extended the analyses we could conduct in blood by examining the PTSD-associated blood *CLDN5* DNAm loci in association with *CLDN5* expression in the three brain regions, covarying for age at death, sex, PMI, 3 PCs, 3 qSVs, sequencing run ID, and RNA-derived brain cell type estimates. We examined *CLDN5* expression in each brain region in association with the 7 types of estimated cell types to test hypothesized associations between *CLDN5* and endothelial cells, covarying for age at death, sex, PMI, 3 PCs, 3 qSVs, and sequencing run ID, and correcting across the cell types and brain regions using FDR.

## 3. Results

### 3.1 Peripheral *CLDN5* DNAm and Current PTSD Severity

Analyses examining associations between current PTSD symptom severity and each of the 19 *CLDN5* probes derived from blood samples revealed 2 nominally significant and an additional 3 FDR-corrected significant associations (Table 2). The associations that surpassed the multiple testing correction were between PTSD severity and cg21872764 (β = .086, *p* = .002, *p-*_adj_ = .032), cg00804505 (β = -.080, *p* = .004, *p-*_adj_ = .032), and cg17411190 (β = .068, *p* = .005, *p-*_adj_ = .032). These three CpG sites were not highly correlated with each other (strongest *r* = .262; Table S1), suggesting that their associations with PTSD severity represented independent effects.

**Table 2.** Associations between Current PTSD Severity and CLDN5 DNA Methylation in Blood (N = 1311)

| Probe | B (unstd) | $\beta$ | $p$ | $p_{\text{adj}}$ |
| --- | --- | --- | --- | --- |
| cg00189989 | .038 | .041 | .144 | .249 |
| **cg00804504** | **-.094** | **-.080** | **.004** | **.032** |
| cg00811132 | .074 | .050 | .061 | .193 |
| cg04463638 | -.029 | -.027 | .334 | .428 |
| cg05460329 | -.045 | -.058 | .038 | .144 |
| cg05498726 | -.028 | -.025 | .360 | .428 |
| cg06315607 | -.017 | -.012 | .660 | .660 |
| cg06340942 | -.055 | -.044 | .110 | .232 |
| cg09092054 | -.036 | -.035 | .187 | .232 |
| cg09446908 | -.074 | -.046 | .103 | .232 |
| cg11450827 | .038 | .046 | .110 | .232 |
| cg13114849 | .025 | .043 | .123 | .234 |
| cg14553765 | -.039 | -.032 | .254 | .371 |
| cg16773741 | -.066 | -.059 | .032 | .144 |
| **cg17411190** | **.098** | **.068** | **.005** | **.032** |
| cg17577122 | -.028 | -.018 | .523 | .585 |
| cg17583256 | .018 | .015 | .582 | .614 |
| cg20486569 | .036 | .022 | .341 | .428 |
| **cg21872764** | **.128** | **.086** | **.002** | **.032** |

*Note.* Covariates included in the model were sex, age, proportional white blood cell types, and the first 3 ancestry principal components. Significant associations are shown in bold font. Unstd = unstandardized; adj = adjusted.

To test the three associations between PTSD severity and the CpG loci for potential confounds, we added lifetime trauma exposure, DNAm smoking score, and MDD diagnosis to each model (each in separate regressions). For the follow-up models involving cg00804505 and cg17411190, none of these covariates were significantly associated with the probe, while PTSD severity remained significantly associated. When adding lifetime trauma exposure to the model involving cg21872764, the association between the probe and PTSD severity was no longer significant (*p* = .137) while trauma exposure was strongly related to DNAm at this locus (β = .109, *p* = .000185). Additional analysis revealed that the association between trauma exposure and cg21872764 (and only this probe) would withstand an FDR correction across all 19 probes (*p_-_*_adj_ = .000589). This association between trauma exposure and the probe also remained significant with the DNAm smoking score and MDD in the model. We examined *CLDN5* candidate SNPs (rs10314 and rs885985) for their associations with the three trauma/PTSD-associated DNAm probes in the EUR cohort (*n* = 873). These variants are 158 to 3254 bp away from the 3 DNAm loci. The variant rs10314 was associated with two of the PTSD-related probes: cg00804504 (*p*_-adj_ = 5.08E-34) and cg17411190 (*p*_-adj_ = 0.000125; Table S2). Similarly, rs885985 was associated with all 3 PTSD-associated probes: cg00804504 (*p*_-adj_ = 5.02E-18), cg17411190 (*p*_-adj_ = 7.69E-18), and cg21872764 (*p*_-adj_ = 0.0000908). Neither SNP was associated with lifetime trauma exposure (*p*s = .094 to .249) nor current PTSD severity (*p*s = .081 to .129), covarying for age, sex, and 3 ancestry substructure PCs. Though the SNPs were associated with the 3 PTSD-associated DNAm probes, PTSD severity and trauma exposure were still significantly associated with their respective DNAm probes with the SNPs included in the models (Table S3).

### 3.2 *CLDN5* DNAm and Biomarkers of Inflammation and Neuropathology

We examined cross-sectional associations between the 3 FDR-significant trauma/PTSD-associated loci and peripheral IL-6, TNFα, GFAP, Aβ-42/Aβ-40 ratio, and NFL (covarying for sex, age, and WBCs). There was an association between the peak probe cg21872764 and NFL (β = .066, *p* = .011, *p*_-adj_ = .033; Table 3). A follow-up cross-sectional mediation path model revealed an indirect association between trauma exposure and NFL via cg21872764 (indirect β = .009, *p* = .015; Figure 1; Supplementary Materials).

**Figure 1.**
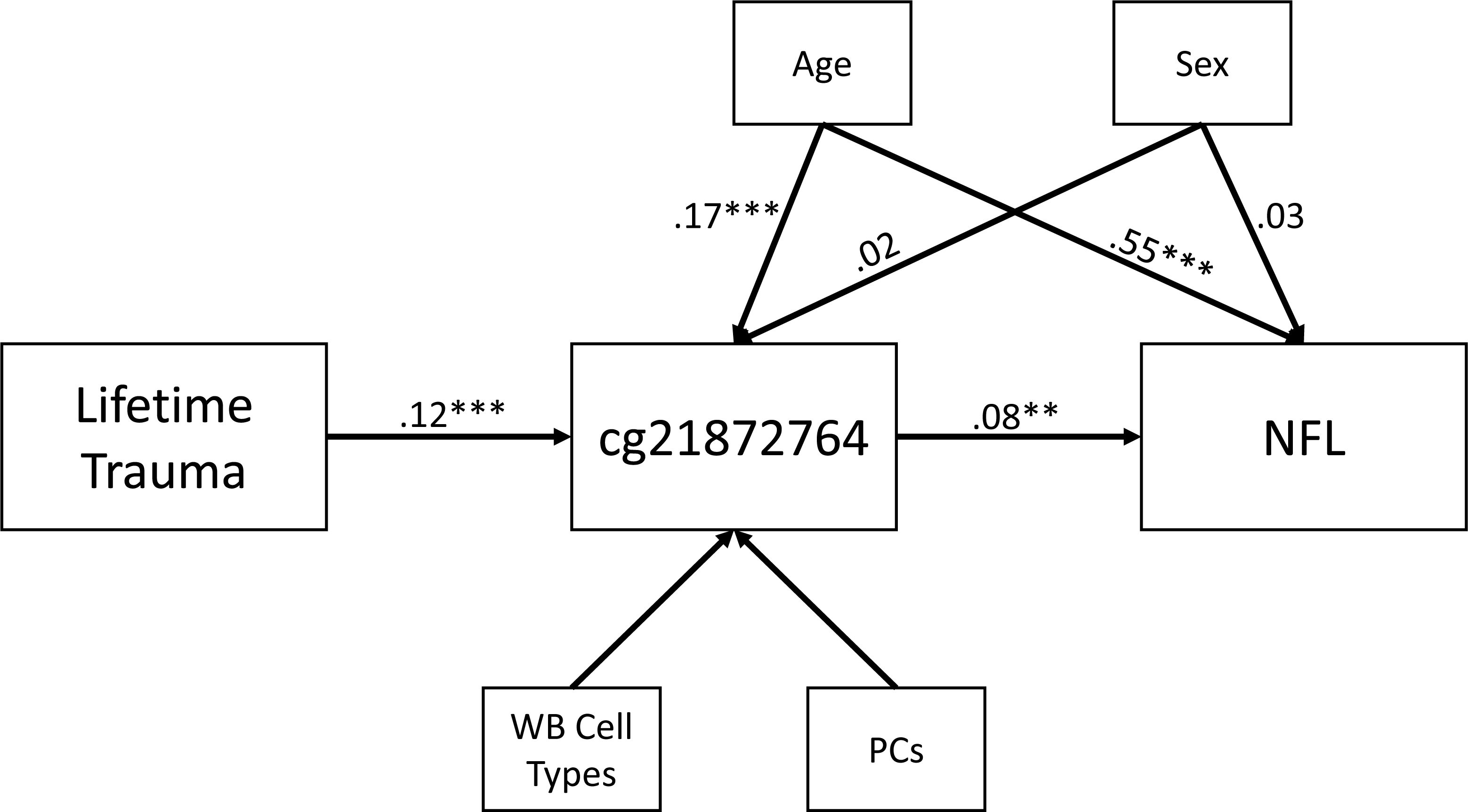
shows the results of the mediation path model in which trauma exposure was indirectly associated with NFL levels via DNAm at cg21872764, covarying for potential demographic and methodological confounds.

**Table 3.** Cross-sectional Associations between Peripheral CLDN5 DNA Methylation and Inflammation and Neuropathology Biomarkers (n = 1097)

| Probe | IL-6 | | TNFα | | GFAP | | Aβ-42/Aβ-40 | | NFL | |
| --- | --- | --- | --- | --- | --- | --- | --- | --- | --- | --- |
| | $\beta$ | *p* | $\beta$ | *p* | $\beta$ | *p* | $\beta$ | *p* | $\beta$ | *p* |
| cg00804504 | -.041 | .160 | -.006 | .843 | -.043 | .128 | .043 | .100 | -.043 | .092 |
| cg17411190 | -.021 | .538 | -.003 | .921 | .058 | .085 | -.040 | .192 | -.012 | .680 |
| cg21872764 | -.022 | .462 | .031 | .296 | -.011 | .721 | .025 | .352 | **.066** | **.011<sup>a</sup>** |

*Note.* Covariates included sex, age, and proportional white blood cells. Significant associations are shown in bold font. IL = interleukin; TNFα = tumor necrosis factor alpha; GFAP = glial fibrillary acidic protein; Aβ = amyloid beta; NFL = neurofilament light.
<sup>a</sup>The FDR corrected *p*-value for this probe (corrected across the 5 biomarkers) was significant at .033.

### 3.3 Follow-up in Postmortem Brain Tissue

Analyses in the brain bank cohort revealed that two of the three *CLDN5* loci that were associated with trauma/PTSD in blood were also associated in brain tissue. PTSD was associated with cg17411190 (B = 0.305, β = 0.369, *p* = 0.005) and cg21872764 (B = 0.308, β = 0.246, *p* = 0.034), both in vmPFC (Table 4). The probes showed the same direction of association in both tissues. For completeness, we examined the remaining 16 *CLDN5* probes on the chip in association with PTSD, but none were significant after FDR correction across the probes and brain regions (Table S4).

**Table 4.** Associations between PTSD Diagnosis and Three CLDN5 DNAm Probes in Postmortem Brain Tissue.

| CPG | dlPFC (*n* = 69) |  |  | vmPFC (*n* = 69) |  |  | Motor Cortex (*n* = 68) |  |  |
| --- | --- | --- | --- | --- | --- | --- | --- | --- | --- |
| | B | $\beta$ | *p* | B | $\beta$ | *p* | B | $\beta$ | *p* |
| cg00804504 | -0.338 | -0.285 | 0.055 | -0.109 | -0.104 | 0.494 | 0.035 | 0.030 | 0.842 |
| **cg17411190** | 0.114 | 0.129 | 0.381 | **0.305** | **0.369** | **0.005** | 0.196 | 0.238 | 0.148 |
| **cg21872764** | 0.243 | 0.196 | 0.137 | **0.308** | **0.246** | **0.034** | 0.144 | 0.129 | 0.383 |

*Note.* Significant associations are shown in bold font. dlPFC = dorsolateral prefrontal cortex; vmPFC = ventromedial prefrontal cortex.

None of the three PTSD-associated DNAm loci were significantly associated with *CLDN5* expression in any brain region. For completeness, we examined all *CLDN5* probes on the chip and found two nominally significant associations with *CLDN5* expression that did not withstand correction for multiple testing: cg09446908 (B = -0.343, β = -0.220, *p* = 0.005) and cg17577122 (B = -0.118, β = -0.174, *p* = 0.028), both in vmPFC (Table S5). Comparison of *CLDN5* expression with estimated cell types confirmed expected associations between *CLDN5* RNA and endothelial cells in all three brain regions (βs = 0.731 - 0.743 and *p*_-adj_ = 7.336×10^-17^ – 4.907×10^-12^; Table 5). Multiple testing adjusted associations also emerged between *CLDN5* expression and estimated mural cells, astrocytes, and microglia in all three regions, neurons in dlPFC and motor cortex, and red blood cells in dlPFC and vmPFC (Table 5). The direction of these associations was positive except for neurons.

**Table 5.** Associations between the CLDN5 Expression and Estimated Cell Type Proportions in Postmortem Brain Tissues.

| Cell Type | dlPFC (n = 93) |  |  |  | vmPFC (n = 86) |  |  |  | Motor cortex (n = 89) |  |  |  |
| --- | --- | --- | --- | --- | --- | --- | --- | --- | --- | --- | --- | --- |
| | B | $\beta$ | *p* | *p*-adj | B | $\beta$ | *p* | *p*-adj | B | $\beta$ | *p* | *p*-adj |
| Astrocyte | **0.570** | **0.566** | **4.424E-08** | **1.548E-07** | **0.218** | **0.247** | **0.025** | **0.033** | **0.368** | **0.326** | **3.539E-04** | **0.001** |
| Endothelial | **0.680** | **0.739** | **3.493E-18** | **7.336E-17** | **0.609** | **0.731** | **7.010E-13** | **4.907E-12** | **0.621** | **0.743** | **1.747E-13** | **1.835E-12** |
| Microglia | **0.702** | **0.584** | **1.085E-08** | **4.557E-08** | **0.500** | **0.410** | **0.001** | **0.001** | **0.451** | **0.363** | **0.002** | **0.004** |
| Mural | **0.551** | **0.625** | **2.730E-11** | **1.433E-10** | **0.397** | **0.505** | **9.667E-08** | **2.900E-07** | **0.349** | **0.495** | **3.592E-07** | **9.430E-07** |
| Neuron | **-0.358** | **-0.413** | **2.988E-06** | **6.973E-06** | -0.154 | -0.152 | 0.174 | 0.202 | **-0.276** | **-0.264** | **0.003** | **0.005** |
| Oligodendrocyte | 0.067 | 0.054 | 0.561 | 0.589 | 0.037 | 0.027 | 0.829 | 0.829 | 0.121 | 0.086 | 0.289 | 0.319 |
| RBC | **0.267** | **0.296** | **0.005** | **0.008** | **0.285** | **0.291** | **0.012** | **0.017** | 0.134 | 0.161 | 0.126 | 0.156 |

*Note.* *p*-adj = adjusted *p*-value via an FDR correction across 21 tests (3 regions \* 7 cell types). Significant associations are shown in bold font. dlPFC = dorsolateral prefrontal cortex; vmPFC = ventromedial prefrontal cortex; RBC = red blood cells.

## 4. Discussion

### 4.1 Overview

In this study, we expanded on the burgeoning literature suggesting a role for *CLDN5* in BBB permeability, depression, and neuroinflammation in stressed organisms (Menard et al., 2017). We found preliminary evidence that the number of different types of traumatic events experienced across the lifetime was associated with *CLDN5* DNAm at cg21872764. Two additional *CLDN5* DNAm loci, cg00804505 and cg17411190, were associated with PTSD severity. Two of these trauma and PTSD-associated probes (the peak probe, cg21872764 and cg17411190) also showed associations with PTSD when methylation was measured in vmPFC, a region previously implicated in postmortem *CLDN5* expression studies of depression (Dion-Albert et al., 2022). This is consistent with evidence for correlations between these loci in blood and brain when measured in frontal and temporal regions in reference databases (e.g., Blood-Brain Epigenetic Concordance or BECon; Edgar et al., 2017). We did not observe associations between PTSD diagnosis and *CLDN5* DNAm in dlPFC or motor cortex. This may suggest the vmPFC is more vulnerable to stress-related alterations in *CLDN5* DNAm. Meta-analyses suggest that PTSD is associated with vmPFC hypoactivity in the context of amygdala hyperactivity (the two regions evidence bidirectional projections between them), consistent with the notion of insufficient inhibition of emotional arousal in PTSD (Hayes et al., 2012). Whether BBB integrity and *CLDN5* play a role in this is unknown and requires additional research.

Associations between trauma and PTSD and the *CLDN5* loci were not better accounted for by rs10314 and rs885985, despite these genotypes showing associations with *CLDN5* DNAm. Results are broadly consistent with the preclinical literature suggesting a role for cldn5 in stress-related depression-like phenotypes (Menard et al., 2017). Our results raise the possibility that *CLDN5* is associated with both the stressor itself (i.e., trauma exposure) and the chronic psychiatric stress response (i.e., PTSD) and suggest the need for future mechanistic research evaluating the role of *CLDN5* epigenetics in linking trauma and PTSD to BBB degradation and related neuropathology.

We found that the peak trauma and PTSD-associated locus (in blood and brain, respectively), cg21872764, evidenced a positive cross-sectional association with peripheral NFL. Additionally, our preliminary cross-sectional mediation model suggested that the association between trauma exposure and plasma NFL was mediated by blood DNAm at cg21872764. Prior preclinical research suggests that BBB permeability is strongly correlated with serum NFL levels at multiple timepoints following experimental head injury (Arena et al., 2002). Thus, to the extent that cg21872764 DNAm in blood is a marker for the same locus in brain (Edgar et al., 2017), DNAm at this locus may signal stress-associated BBB disruption. Analysis of data from brain tissue also revealed that *CLDN5* expression was associated with cell type estimates in each brain region, with the most significant and consistent effects evident for endothelial cells, which were positively associated with *CLDN5* expression. We observed nominally significant associations between *CLDN5* probes cg09446908 and cg17577122 (that were not associated with trauma or PTSD in blood or brain) and reduced *CLND5* expression in vmPFC. One possible explanation for this pattern of results is that trauma and PTSD-associated alterations in *CLDN5* DNAm give rise to *CLDN5* downregulation in endothelial cells, allowing for central biomarkers of neuropathology, like NFL, to cross the BBB to the periphery. Consistent with this possibility, DNAm at two *CLDN5* loci that were nominally associated with PTSD severity in blood in this study, cg05460329 and cg16773741, were previously found to be associated with worse cognitive performance over time (at the epigenome-wide level of statistical significance) when measured in dlPFC (Hüls et al., 2022). As these data are cross-sectional, we can make no causal or directional claims, however, the evidence for relationships between PTSD, *CLDN5* DNAm, and biomarkers of neuropathology highlight the need for future studies to test this.

### 4.2 Future Research

Additional research is needed to test the replicability of these associations, determine their overlap with other stress-related psychiatric conditions like depression, and evaluate temporal associations between PTSD, *CLDN5* DNAm, and neuropathology biomarkers. *CLDN5* epigenetics may be a useful target for future intervention research aimed at improving BBB integrity and reducing psychological symptoms. The antidepressants imipramine and fluoxetine (Menard et al., 2017; Sun et al., 2024) and the mood stabilizer lithium (Taler et al., 2020) appear to modulate cldn5 expression, reduce stress-associated depressive phenotypes, and protect the brain from inflammatory insults in rodents. Similarly, experimental agents, such as a glycogen synthase kinase-3 inhibitor (Cheng et al., 2018) have been shown to reduce depression-like symptoms and alter cldn5 expression in rodent brains.

Nonpharmacological interventions may also influence *CLDN5* expression. A recent study found that positive environmental conditions (an enriched environment) mitigated the effects of early life stress (maternal separation) on cldn5 expression, BBB permeability, and depressive-like behaviors in mice (Ansari et al., 2025). Collectively, these studies suggest that *CLDN5* epigenetics may be altered through environmental or pharmacological approaches, with concomitant improvement in BBB integrity and symptom reduction. Future research could evaluate if any of these interventions operating on *CLDN5* also reduce future neurological risk among those with PTSD.

### 4.3 Study Limitations

The results of this study should be considered preliminary in light of several limitations. The study was comprised of primarily male Veterans with PTSD and may not generalize to non-Veterans or women. Further, the relatively young mean age of the TRACTS subset (in their early 30s) and the exclusion of Veterans with cognitive or neurological disease (other than that related to TBI) from that cohort may have limited the range of NFL in plasma and made it more difficult to observe associations with *CLDN5* DNAm. This concern is offset to an extent by the inclusion of the older NCPTSD cohort (57% of the sample) without these exclusions. Still, follow-up studies in older Veterans with greater neurological risk are needed to more comprehensively evaluate *CLDN5* DNAm in association with PTSD and neurological impairment. The brain bank cohort was small and power was limited in these analyses.

We only had access to data from three brain regions and it is possible that associations between *CLDN5* DNAm and expression may differ in other regions. The differential associations observed across brain regions require replication in larger samples. We did not have a measure of trauma exposure in the brain bank cohort that would allow us to disentangle its associations with *CLDN5* DNAm apart from those of PTSD. We also could not cleanly separate effects related to PTSD from those associated with MDD in the brain bank because the majority of PTSD cases had comorbid MDD. Brain bank results were broadly consistent with those from our living cohort (in which we were able to covary for MDD), helping to mitigate this concern. We did not have data concerning CLDN5 protein levels in blood or brain, BBB staining metrics, or BBB permeability neuroimaging (e.g., via positron emission tomography) that would have allowed for more direct examination of BBB permeability. Most importantly, we cannot infer causality or temporality from these cross-sectional associations and additional research is needed to address such questions.

### 4.4 Conclusions

In this first-ever study of associations between traumatic stress and a gene critically responsible for tight junction integrity of the BBB, we found evidence for cross-sectional associations between trauma exposure, PTSD, and *CLDN5* DNAm and between *CLDN5* DNAm and markers of neuropathology. We suspect that chronic PTSD symptoms function as a stressor that may alter *CLDN5* DNAm, leading to decreased *CLDN5* expression in brain endothelial cells and increased BBB permeability, allowing for bidirectional passage of neuroinflammatory molecules across the BBB. These preliminary cross-sectional results may help to explain the link between traumatic stress and neuropathology and highlight the role of DNAm in sensitivity to environmental insult. If these results are replicated and future studies support a mechanistic role for *CLDN5*, this could inform the development of novel therapeutic approaches for reducing risk for neurological disease among those with PTSD.

## Data Availability

The data that this manuscript is based on are held in VA data repositories. Qualified investigators may write to the first author to obtain detailed information on the process of applying to access data from a data repository.

## Acknowledgements

This work was supported in party by Merit Review Award Number I01 CX-001276-01 from the United States (U.S.) Department of Veterans Affairs Clinical Sciences R&D (CSRD) Service and by 2 I50RX003001-06 from the U.S. Department of Veterans Affairs, Rehabilitation Research and Development Program. Research reported in this publication was supported by the National Institute On Aging of the National Institutes of Health under Award Number RF1AG068121/4R01AG068121-02 and by National Institute On Aging of the National Institutes of Health under Award Number 1R21AG061367-01. The content is solely the responsibility of the authors and does not necessarily represent the official views of the National Institutes of Health, the U.S. Department of Veterans Affairs, or the United States Government.

The Traumatic Stress Brain Research Group is comprised of the following individuals: Victor E. Alvarez, David Benedek, Alicia Che, Dianne A. Cruz, David A. Davis, Matthew J. Girgenti, Ellen Hoffman, Paul E. Holtzheimer, Alfred Kaye, John H. Krystal, Adam T. Labadorf, Terence M. Keane, Ann McKee, Brian Marx, Crystal Noller, Meghan Pierce, William K. Scott, Paula Schnurr, Krista DiSano, Thor Stein, Douglas E. Williamson, Keith A. Young.

## Disclosures

All authors report no financial or other conflicts of interest.

## Notes

### Competing Interest Statement

The authors have declared no competing interest.

### Author Declarations

The IRB of the VA Boston Healthcare System gave ethical approval for this work.

## References

Arena, J. D., Smith, D. H., Diaz Arrastia, R., Cullen, D. K., Xiao, R., Fan, J., Harris, D. C., Lynch, C. E., & Johnson, V. E. (2024). The neuropathological basis of elevated serum neurofilament light following experimental concussion. Acta Neuropathologica Communications, 12(1), 189. 10.1186/s40478-024-01883-z

Benjamini Y. & Hochberg Y. (1995). Controlling the false discovery rate: A practical and powerful approach to multiple testing. Journal of the Royal Statistical Society: Series B (Methodological), 57, 289–300.

Blake, D. D., Weathers, F. W., Nagy, L. M., Kaloupek, D. G., Gusman, F. D., Charney, D. S., & Keane, T. M. (1995). The development of a Clinician-Administered PTSD Scale. Journal of Traumatic Stress, 8(1), 75–90. 10.1007/BF02105408

Bourassa, K. J., & Sbarra, D. A. (2024). Trauma, adversity, and biological aging: Behavioral mechanisms relevant to treatment and theory. Translational Psychiatry, 14(1), 285. 10.1038/s41398-024-03004-9

Camire, R. B., Beaulac, H. J., & Willis, C. L. (2015). Transitory loss of glia and the subsequent modulation in inflammatory cytokines/chemokines regulate paracellular claudin-5 expression in endothelial cells. Journal of Neuroimmunology, 284, 57–66. 10.1016/j.jneuroim.2015.05.008

Cheng, Y., Desse, S., Martinez, A., Worthen, R. J., Jope, R. S., & Beurel, E. (2018). TNFα disrupts blood brain barrier integrity to maintain prolonged depressive-like behavior in mice. Brain, Behavior, and Immunity, 69, 556–567. 10.1016/j.bbi.2018.02.003

Christensen, B. C., Houseman, E. A., Marsit, C. J., Zheng, S., Wrensch, M. R., Wiemels, J. L., Nelson, H. H., Karagas, M. R., Padbury, J. F., Bueno, R., Sugarbaker, D. J., Yeh, R. F., Wiencke, J. K., & Kelsey, K. T. (2009). Aging and environmental exposures alter tissue-specific DNA methylation dependent upon CpG island context. PLoS Genetics, 5(8), e1000602. 10.1371/journal.pgen.1000602

Dalvie, S., Chatzinakos, C., Al Zoubi, O., Georgiadis, F., PGC-PTSD Systems Biology workgroup, Lancashire, L., & Daskalakis, N. P. (2021). From genetics to systems biology of stress-related mental disorders. Neurobiology of Stress, 15, 100393. 10.1016/j.ynstr.2021.100393

Dion-Albert, L., Cadoret, A., Doney, E., Kaufmann, F. N., Dudek, K. A., Daigle, B., Parise, L. F., Cathomas, F., Samba, N., Hudson, N., Lebel, M., Signature Consortium, Campbell, M., Turecki, G., Mechawar, N., & Menard, C. (2022). Vascular and blood-brain barrier-related changes underlie stress responses and resilience in female mice and depression in human tissue. Nature Communications, 13(1), 164. 10.1038/s41467-021-27604-x

Edgar, R. D., Jones, M. J., Meaney, M. J., Turecki, G., & Kobor, M. S. (2017). BECon: a tool for interpreting DNA methylation findings from blood in the context of brain. Translational Psychiatry, 7(8), e1187. 10.1038/tp.2017.171

First, M. B., Spitzer, R., Gibbon, M., & Williams, J. (1994). Structured clinical interview for axis I DSM-IV disorders—patient edition (SCID-I/P, version 2.0). Biometrics Research Department, New York State Psychiatric Institute.

First, M. B., Williams, J. B. W., Karg, R. S., & Spitzer, R. L. (2015). Structured clinical interview for DSM-5—research version (SCID-5 for DSM-5, Research Version; SCID-5-RV). American Psychiatric Association, Arlington, VA.

Friedman, M. J., Huber, B. R., Brady, C. B., Ursano, R. J., Benedek, D. M., Kowall, N. W., McKee, A. C., & Traumatic Stress Brain Research Group (2017). VA’s National PTSD Brain Bank: A national resource for research. Current Psychiatry Reports, 19(10), 73. 10.1007/s11920-017-0822-6

Friis, T., Wikström, A. K., Acurio, J., León, J., Zetterberg, H., Blennow, K., Nelander, M., Åkerud, H., Kaihola, H., Cluver, C., Troncoso, F., Torres-Vergara, P., Escudero, C., & Bergman, L. (2022). Cerebral biomarkers and blood-brain barrier integrity in preeclampsia. Cells, 11(5), 789. 10.3390/cells11050789

Gal, Z., Torok, D., Gonda, X., Eszlari, N., Anderson, I. M., Deakin, B., Juhasz, G., Bagdy, G., & Petschner, P. (2023). Inflammation and blood-brain barrier in depression: Interaction of CLDN5 and IL6 gene variants in stress-induced depression. The International Journal of Neuropsychopharmacology, 26(3), 189–197. 10.1093/ijnp/pyac079

Gonzales, M. M., Wiedner, C., Wang, C. P., Liu, Q., Bis, J. C., Li, Z., Himali, J. J., Ghosh, S., Thomas, E. A., Parent, D. M., Kautz, T. F., Pase, M. P., Aparicio, H. J., Djoussé, L., Mukamal, K. J., Psaty, B. M., Longstreth, W. T., Jr, Mosley, T. H., Jr, Gudnason, V., Mbangdadji, D., … Satizabal, C. L. (2022). A population-based meta-analysis of circulating GFAP for cognition and dementia risk. Annals of Clinical and Translational Neurology, 9(10), 1574–1585. 10.1002/acn3.51652

Greene, C., Hanley, N., & Campbell, M. (2019). Claudin-5: Gatekeeper of neurological function. Fluids and Barriers of the CNS, 16(1), 3. 10.1186/s12987-019-0123-z

Greene, C., Hanley, N., & Campbell, M. (2020). Blood-brain barrier associated tight junction disruption is a hallmark feature of major psychiatric disorders. Translational Psychiatry, 10(1), 373. 10.1038/s41398-020-01054-3

Greene, C., Kealy, J., Humphries, M. M., Gong, Y., Hou, J., Hudson, N., Cassidy, L. M., Martiniano, R., Shashi, V., Hooper, S. R., Grant, G. A., Kenna, P. F., Norris, K., Callaghan, C. K., Islam, M. D., O’Mara, S. M., Najda, Z., Campbell, S. G., Pachter, J. S., Thomas, J., … Campbell, M. (2018). Dose-dependent expression of claudin-5 is a modifying factor in schizophrenia. Molecular Psychiatry, 23(11), 2156–2166. 10.1038/mp.2017.156

Guintivano, J., Aryee, M. J., & Kaminsky, Z. A. (2013). A cell epigenotype specific model for the correction of brain cellular heterogeneity bias and its application to age, brain region and major depression. Epigenetics, 8(3), 290–302. 10.4161/epi.23924

Hagenauer, M. H., Schulmann, A., Li, J. Z., Vawter, M. P., Walsh, D. M., Thompson, R. C., Turner, C. A., Bunney, W. E., Myers, R. M., Barchas, J. D., Schatzberg, A. F., Watson, S. J., & Akil, H. (2018). Inference of cell type content from human brain transcriptomic datasets illuminates the effects of age, manner of death, dissection, and psychiatric diagnosis. PloS One, 13(7), e0200003. 10.1371/journal.pone.0200003

Hashimoto, Y., Campbell, M., Tachibana, K., Okada, Y., & Kondoh, M. (2021). Claudin-5: A pharmacological target to modify the permeability of the blood-brain barrier. Biological & Pharmaceutical Bulletin, 44(10), 1380–1390. 10.1248/bpb.b21-00408

Hashimoto, Y., Greene, C., Munnich, A., & Campbell, M. (2023). The CLDN5 gene at the blood-brain barrier in health and disease. Fluids and Barriers of the CNS, 20(1), 22. 10.1186/s12987-023-00424-5

Hüls, A., Robins, C., Conneely, K. N., Edgar, R., De Jager, P. L., Bennett, D. A., Wingo, A. P., Epstein, M. P., & Wingo, T. S. (2022). Brain DNA methylation patterns in CLDN5 associated with cognitive decline. Biological Psychiatry, 91(4), 389–398. 10.1016/j.biopsych.2021.01.015

Jack, C. R., Jr, Andrews, S. J., Beach, T. G., Buracchio, T., Dunn, B., Graf, A., Hansson, O., Ho, C., Jagust, W., McDade, E., Molinuevo, J. L., Okonkwo, O. C., Pani, L., Rafii, M. S., Scheltens, P., Siemers, E., Snyder, H. M., Sperling, R., Teunissen, C. E., & Carrillo, M. C. (2024). Revised criteria for the diagnosis and staging of Alzheimer’s disease. Nature Medicine, 30(8), 2121–2124. 10.1038/s41591-024-02988-7

Kealy, J., Greene, C., & Campbell, M. (2020). Blood-brain barrier regulation in psychiatric disorders. Neuroscience Letters, 726, 133664. 10.1016/j.neulet.2018.06.033

Keaney, J., Walsh, D. M., O’Malley, T., Hudson, N., Crosbie, D. E., Loftus, T., Sheehan, F., McDaid, J., Humphries, M. M., Callanan, J. J., Brett, F. M., Farrell, M. A., Humphries, P., & Campbell, M. (2015). Autoregulated paracellular clearance of amyloid-β across the blood-brain barrier. Science Advances, 1(8), e1500472. 10.1126/sciadv.1500472

Kim, K. Y., Shin, K. Y., & Chang, K. A. (2023). GFAP as a potential biomarker for Alzheimer’s disease: A systematic review and meta-analysis. Cells, 12(9), 1309. 10.3390/cells12091309

Koestler, D. C., Jones, M. J., Usset, J., Christensen, B. C., Butler, R. A., Kobor, M. S., Wiencke, J. K., & Kelsey, K. T. (2016). Improving cell mixture deconvolution by identifying optimal DNA methylation libraries (IDOL). BMC Bioinformatics, 17, 120. 10.1186/s12859-016-0943-7

Kubany, E. S., Haynes, S. N., Leisen, M. B., Owens, J. A., Kaplan, A. S., Watson, S. B., & Burns, K. (2000). Development and preliminary validation of a brief broad-spectrum measure of trauma exposure: The Traumatic Life Events Questionnaire. Psychological Assessment, 12(2), 210–224. 10.1037//1040-3590.12.2.210

Leek, J. T., Johnson, W. E., Parker, H. S., Jaffe, A. E., & Storey, J. D. (2012). The sva package for removing batch effects and other unwanted variation in high-throughput experiments. *Bioinformatics (Oxford*, England), 28(6), 882–883. 10.1093/bioinformatics/bts034

Lewczuk, P., Esselmann, H., Otto, M., Maler, J. M., Henkel, A. W., Henkel, M. K., Eikenberg, O., Antz, C., Krause, W. R., Reulbach, U., Kornhuber, J., & Wiltfang, J. (2004). Neurochemical diagnosis of Alzheimer’s dementia by CSF Abeta42, Abeta42/Abeta40 ratio and total tau. Neurobiology of Aging, 25(3), 273–281. 10.1016/S0197-4580(03)00086-1

Li, S., Wong, E. M., Bui, M., Nguyen, T. L., Joo, J. E., Stone, J., Dite, G. S., Giles, G. G., Saffery, R., Southey, M. C., & Hopper, J. L. (2018). Causal effect of smoking on DNA methylation in peripheral blood: a twin and family study. Clinical Epigenetics, 10, 18. 10.1186/s13148-018-0452-9

Li, Z., Zhang, J., Halbgebauer, S., Chandrasekar, A., Rehman, R., Ludolph, A., Boeckers, T., Huber-Lang, M., Otto, M., Roselli, F., & Heuvel, F. O. (2021). Differential effect of ethanol intoxication on peripheral markers of cerebral injury in murine blunt traumatic brain injury. Burns & Trauma, 9, tkab027. 10.1093/burnst/tkab027

Logue, M. W., Baldwin, C., Guffanti, G., Melista, E., Wolf, E. J., Reardon, A. F., Uddin, M., Wildman, D., Galea, S., Koenen, K. C., & Miller, M. W. (2013). A genome-wide association study of post-traumatic stress disorder identifies the retinoid-related orphan receptor alpha (RORA) gene as a significant risk locus. Molecular Psychiatry, 18(8), 937–942. 10.1038/mp.2012.113

Logue, M. W., Miller, M. W., Sherva, R., Zhang, R., Harrington, K. M., Fonda, J. R., Merritt, V. C., Panizzon, M. S., Hauger, R. L., Wolf, E. J., Neale, Z., Gaziano, J. M., & Million Veteran Program (2023). Alzheimer’s disease and related dementias among aging veterans: Examining gene-by-environment interactions with post-traumatic stress disorder and traumatic brain injury. Alzheimer’s & Dementia, 19(6), 2549–2559. 10.1002/alz.12870

Logue, M. W., Miller, M. W., Wolf, E. J., Huber, B. R., Morrison, F. G., Zhou, Z., Zheng, Y., Smith, A. K., Daskalakis, N. P., Ratanatharathorn, A., Uddin, M., Nievergelt, C. M., Ashley-Koch, A. E., Baker, D. G., Beckham, J. C., Garrett, M. E., Boks, M. P., Geuze, E., Grant, G. A., Hauser, M. A., … Traumatic Stress Brain Study Group (2020). An epigenome-wide association study of posttraumatic stress disorder in US veterans implicates several new DNA methylation loci. Clinical Epigenetics, 12(1), 46. 10.1186/s13148-020-0820-0

Love, M. I., Huber, W., & Anders, S. (2014). Moderated estimation of fold change and dispersion for RNA-seq data with DESeq2. Genome Biology, 15(12), 550. 10.1186/s13059-014-0550-8

McConnell, H. L., & Mishra, A. (2022). Cells of the blood-brain barrier: An overview of the neurovascular unit in health and disease. *Methods in Molecular Biology (Clifton*, N.J.), 2492, 3–24. 10.1007/978-1-0716-2289-6_1

McGlinchey, R. E., Milberg, W. P., Fonda, J. R., & Fortier, C. B. (2017). A methodology for assessing deployment trauma and its consequences in OEF/OIF/OND veterans: The TRACTS longitudinal prospective cohort study. International Journal of Methods in Psychiatric Research, 26(3), e1556. 10.1002/mpr.1556

Medina-Rodriguez, E. M., & Beurel, E. (2022). Blood brain barrier and inflammation in depression. Neurobiology of Disease, 175, 105926. 10.1016/j.nbd.2022.105926

Menard, C., Pfau, M. L., Hodes, G. E., Kana, V., Wang, V. X., Bouchard, S., Takahashi, A., Flanigan, M. E., Aleyasin, H., LeClair, K. B., Janssen, W. G., Labonté, B., Parise, E. M., Lorsch, Z. S., Golden, S. A., Heshmati, M., Tamminga, C., Turecki, G., Campbell, M., Fayad, Z. A., … Russo, S. J. (2017). Social stress induces neurovascular pathology promoting depression. Nature Neuroscience, 20(12), 1752–1760. 10.1038/s41593-017-0010-3

Mighdoll, M. I., Deep-Soboslay, A., Bharadwaj, R. A., Cotoia, J. A., Benedek, D. M., Hyde, T. M., & Kleinman, J. E. (2018). Implementation and clinical characteristics of a posttraumatic stress disorder brain collection. Journal of Neuroscience Research, 96(1), 16–20. 10.1002/jnr.24093

Miller, M. W., & Sadeh, N. (2014). Traumatic stress, oxidative stress and post-traumatic stress disorder: neurodegeneration and the accelerated-aging hypothesis. Molecular Psychiatry, 19(11), 1156–1162. 10.1038/mp.2014.111

Miller, M. W., Lin, A. P., Wolf, E. J., & Miller, D. R. (2018). Oxidative stress, inflammation, and neuroprogression in chronic PTSD. Harvard Review of Psychiatry, 26(2), 57–69. 10.1097/HRP.0000000000000167

Miller, M. W., Wolf, E. J., Zhao, X., Logue, M. W., & Hawn, S. E. (2024). An EWAS of dementia biomarkers and their associations with age, African ancestry, and PTSD. Clinical Epigenetics, 16(1), 38. 10.1186/s13148-024-01649-3

Ni, K., Zhu, J., Xu, X., Liu, Y., Yang, S., Huang, Y., Xu, R., Jiang, L., Zhang, J., Zhang, W., & Ma, Z. (2022). Hippocampal activated microglia may contribute to blood-brain barrier impairment and cognitive dysfunction in post-traumatic stress disorder-like rats. Journal of Molecular Neuroscience: MN, 72(5), 975–982. 10.1007/s12031-022-01981-4

Nievergelt, C. M., Maihofer, A. X., Atkinson, E. G., Chen, C. Y., Choi, K. W., Coleman, J. R. I., Daskalakis, N. P., Duncan, L. E., Polimanti, R., Aaronson, C., Amstadter, A. B., Andersen, S. B., Andreassen, O. A., Arbisi, P. A., Ashley-Koch, A. E., Austin, S. B., Avdibegoviç, E., Babić, D., Bacanu, S. A., Baker, D. G., … Koenen, K. C. (2024). Genome-wide association analyses identify 95 risk loci and provide insights into the neurobiology of post-traumatic stress disorder. Nature Genetics, 56(5), 792–808. 10.1038/s41588-024-01707-9

Nievergelt, C. M., Maihofer, A. X., Klengel, T., Atkinson, E. G., Chen, C. Y., Choi, K. W., Coleman, J. R. I., Dalvie, S., Duncan, L. E., Gelernter, J., Levey, D. F., Logue, M. W., Polimanti, R., Provost, A. C., Ratanatharathorn, A., Stein, M. B., Torres, K., Aiello, A. E., Almli, L. M., Amstadter, A. B., … Koenen, K. C. (2019). International meta-analysis of PTSD genome-wide association studies identifies sex- and ancestry-specific genetic risk loci. Nature Communications, 10(1), 4558. 10.1038/s41467-019-12576-w

Nitta, T., Hata, M., Gotoh, S., Seo, Y., Sasaki, H., Hashimoto, N., Furuse, M., & Tsukita, S. (2003). Size-selective loosening of the blood-brain barrier in claudin-5-deficient mice. The Journal of Cell Biology, 161(3), 653–660. 10.1083/jcb.200302070

Pardridge W. M. (2005). The blood-brain barrier: Bottleneck in brain drug development. NeuroRx: The Journal of the American Society for Experimental NeuroTherapeutics, 2(1), 3–14. 10.1602/neurorx.2.1.3

Peruzzolo, T. L., Pinto, J. V., Roza, T. H., Shintani, A. O., Anzolin, A. P., Gnielka, V., Kohmann, A. M., Marin, A. S., Lorenzon, V. R., Brunoni, A. R., Kapczinski, F., & Passos, I. C. (2022). Inflammatory and oxidative stress markers in post-traumatic stress disorder: A systematic review and meta-analysis. Molecular Psychiatry, 27(8), 3150–3163. 10.1038/s41380-022-01564-0

Ratanatharathorn, A., Boks, M. P., Maihofer, A. X., Aiello, A. E., Amstadter, A. B., Ashley-Koch, A. E., Baker, D. G., Beckham, J. C., Bromet, E., Dennis, M., Garrett, M. E., Geuze, E., Guffanti, G., Hauser, M. A., Kilaru, V., Kimbrel, N. A., Koenen, K. C., Kuan, P. F., Logue, M. W., Luft, B. J., … Smith, A. K. (2017). Epigenome-wide association of PTSD from heterogeneous cohorts with a common multi-site analysis pipeline. American Journal of Medical Genetics. Part B, Neuropsychiatric Genetics: The Official Publication of the International Society of Psychiatric Genetics, 174(6), 619–630. 10.1002/ajmg.b.32568

Ringwald, W. R., Forbes, M. K., & Wright, A. G. C. (2023). Meta-analysis of structural evidence for the Hierarchical Taxonomy of Psychopathology (HiTOP) model. Psychological Medicine, 53(2), 533–546. 10.1017/S0033291721001902

Roberts, A. L., Liu, J., Lawn, R. B., Jha, S. C., Sumner, J. A., Kang, J. H., Rimm, E. B., Grodstein, F., Kubzansky, L. D., Chibnik, L. B., & Koenen, K. C. (2022). Association of posttraumatic stress disorder with accelerated cognitive decline in middle-aged women. JAMA Network Open, 5(6), e2217698. 10.1001/jamanetworkopen.2022.17698

Sántha, P., Veszelka, S., Hoyk, Z., Mészáros, M., Walter, F. R., Tóth, A. E., Kiss, L., Kincses, A., Oláh, Z., Seprényi, G., Rákhely, G., Dér, A., Pákáski, M., Kálmán, J., Kittel, Á., & Deli, M. A. (2016). Restraint stress-induced morphological changes at the blood-brain barrier in adult rats. Frontiers in Molecular Neuroscience, 8, 88. 10.3389/fnmol.2015.00088

Sugden, K., Caspi, A., Elliott, M. L., Bourassa, K. J., Chamarti, K., Corcoran, D. L., Hariri, A. R., Houts, R. M., Kothari, M., Kritchevsky, S., Kuchel, G. A., Mill, J. S., Williams, B. S., Belsky, D. W., Moffitt, T. E., & Alzheimer’s Disease Neuroimaging Initiative* (2022). Association of pace of aging measured by blood-based DNA methylation with age-related cognitive impairment and dementia. Neurology, 99(13), e1402–e1413. 10.1212/WNL.0000000000200898

Sun, Z. W., Wang, X., Zhao, Y., Sun, Z. X., Wu, Y. H., Hu, H., Zhang, L., Wang, S. D., Li, F., Wei, A. J., Feng, H., Xie, F., & Qian, L. J. (2024). Blood-brain barrier dysfunction mediated by the EZH2-Claudin-5 axis drives stress-induced TNF-α infiltration and depression-like behaviors. *Brain*, Behavior, and Immunity, 115, 143–156. 10.1016/j.bbi.2023.10.010

Sun, Z. Y., Wei, J., Xie, L., Shen, Y., Liu, S. Z., Ju, G. Z., Shi, J. P., Yu, Y. Q., Zhang, X., Xu, Q., & Hemmings, G. P. (2004). The CLDN5 locus may be involved in the vulnerability to schizophrenia. European Psychiatry: The Journal of the Association of European Psychiatrists, 19(6), 354–357. 10.1016/j.eurpsy.2004.06.007

Taler, M., Aronovich, R., Henry Hornfeld, S., Dar, S., Sasson, E., Weizman, A., & Hochman, E. (2021). Regulatory effect of lithium on hippocampal blood-brain barrier integrity in a rat model of depressive-like behavior. Bipolar Disorders, 23(1), 55–65. 10.1111/bdi.12962

Teschendorff, A. E., Breeze, C. E., Zheng, S. C., & Beck, S. (2017). A comparison of reference-based algorithms for correcting cell-type heterogeneity in Epigenome-Wide Association Studies. BMC Bioinformatics, 18(1), 105. 10.1186/s12859-017-1511-5

Tobi, E. W., Lumey, L. H., Talens, R. P., Kremer, D., Putter, H., Stein, A. D., Slagboom, P. E., & Heijmans, B. T. (2009). DNA methylation differences after exposure to prenatal famine are common and timing- and sex-specific. Human Molecular Genetics, 18(21), 4046–4053. 10.1093/hmg/ddp353

Vázquez-Liébanas, E., Mocci, G., Li, W., Laviña, B., Reddy, A., O’Connor, C., Hudson, N., Elbeck, Z., Nikoloudis, I., Gaengel, K., Vanlandewijck, M., Campbell, M., Betsholtz, C., & Mäe, M. A. (2024). Mosaic deletion of claudin-5 reveals rapid non-cell-autonomous consequences of blood-brain barrier leakage. Cell Reports, 43(3), 113911. 10.1016/j.celrep.2024.113911

Weathers, F. W., Bovin, M. J., Lee, D. J., Sloan, D. M., Schnurr, P. P., Kaloupek, D. G., Keane, T. M., & Marx, B. P. (2018). The Clinician-Administered PTSD Scale for DSM-5 (CAPS-5): Development and initial psychometric evaluation in military veterans. Psychological Assessment, 30(3), 383–395. 10.1037/pas0000486

Wolf, E. J., Miller, M. W., Hawn, S. E., Zhao, X., Wallander, S. E., McCormick, B., Govan, C., Rasmusson, A., Stone, A., Schichman, S. A., & Logue, M. W. (2024). Longitudinal study of traumatic-stress related cellular and cognitive aging. *Brain*, Behavior, and Immunity, 115, 494–504. 10.1016/j.bbi.2023.11.009

Wolf, E. J., Zhao, X., Hawn, S. E., Morrison, F. G., Zhou, Z., Fein-Schaffer, D., Huber, B., Traumatic Stress Brain Research Group, Miller, M. W., & Logue, M. W. (2021). Gene expression correlates of advanced epigenetic age and psychopathology in postmortem cortical tissue. Neurobiology of Stress, 15, 100371. 10.1016/j.ynstr.2021.100371

Wu, S., Yin, Y., & Du, L. (2022). Blood-brain barrier dysfunction in the pathogenesis of major depressive disorder. Cellular and Molecular Neurobiology, 42(8), 2571–2591. 10.1007/s10571-021-01153-9

Yaffe, K., Vittinghoff, E., Lindquist, K., Barnes, D., Covinsky, K. E., Neylan, T., Kluse, M., & Marmar, C. (2010). Posttraumatic stress disorder and risk of dementia among US veterans. Archives of General Psychiatry, 67(6), 608–613. 10.1001/archgenpsychiatry.2010.61

Yuan, A., & Nixon, R. A. (2021). Neurofilament proteins as biomarkers to monitor neurological diseases and the efficacy of therapies. Frontiers in Neuroscience, 15, 689938. 10.3389/fnins.2021.689938

Zhao, X., Logue, M. W., Hawn, S. E., Neale, Z. E., Zhou, Z., Huber, B. R., Traumatic Stress Brain Research Group, Miller, M. W., & Wolf, E. J. (2022). PTSD, major depression, and advanced transcriptomic age in brain tissue. Depression and Anxiety, 39(12), 824–834. 10.1002/da.23289

