## Supplementary Materials for "Trauma Exposure, PTSD, and Methylation of the Blood Brain Barrier Claudin-5 Gene"

Erika J. Wolf,^1,2^ Xiang Zhao,^3^ Annelise Madison,^1,4,5^ Jack Carbaugh,^6^ Catherine B. Fortier,^6,7,8^

William P. Milberg,^6,7,8^ Traumatic Stress Brain Research Group, Mark W. Logue^1,2,3^ and Mark W. Miller^1,2^

^1^National Center for PTSD at VA Boston Healthcare System, Boston, MA, USA

^2^Boston University Chobanian & Avedisian School of Medicine, Department of Psychiatry, Boston, MA, USA

^3^Boston University School of Public Health, Department of Biostatistics, Boston, MA, USA

^4^VA Boston Healthcare System, Psychology Service, Boston, MA, USA

^5^University of Michigan, Department of Psychology, Ann Arbor, MI

^6^Translational Research Center for TBI and Stress Disorders (TRACTS), VA Boston Healthcare System, Boston, MA, USA

^7^Geriatric Research, Educational and Clinical Center (GRECC), VA Boston Healthcare System, Boston, MA, USA.

^8^Department of Psychiatry, Harvard Medical School, Boston, MA, USA

**DNA and DNA Methylation (DNAm) Methods**

Genome-wide genotype imputation was conducted using the RICOPILI (Lam et al., 2020) procedures based on the Thousand Genomes Phase 3 reference panel (1000 Genomes Project Consortium et al., 2015) in hg19/ GRCh37 (Church et al., 2011).

To process the DNAm data in both blood and brain, we followed procedures developed by the PGC PTSD Epigenetics Working Group (Ratanatharathorn et al., 2017; and its updated version at <https://github.com/PGC-PTSD-EWAS/EPIC_QC>). Briefly, we excluded probes that failed the Illumina type control metrics (https://support.illumina.com/content/dam/illumina-support/documents/documentation/chemistry_documentation/infinium_assays/infinium_hd_methylation/beadarray-controls-reporter-user-guide-1000000004009-00.pdf) or identified as cross-hybridized sites. Probes with a detection p-value > 0.01 were set to missing. Noob background correction was applied using the minfi package (Aryee et al., 2014) to normalize the probes. Methylation-predicted sex was checked against the genotyped sex if available, or the self-reported sex, and mismatches were eliminated. Samples with >10% missing data or that were outside the bounds of the probe intensity threshold (< 50% of the experiment-wide mean or with intensity <2,000 arbitrary units) were excluded. A ComBat adjustment was then implemented for removal of chip and position effects while preserving variation of age, sex, and PTSD diagnosis using the Bioconductor package sva (Leek, Johnson, Parker, Jaffe, & Storey, 2012). Imputation was carried out using k-nearest neighbor method via Bioconductor impute package (Hastie, Tibshirani, Narasimhan, & Chu, 2023).

**Candidate *CLDN5* Genotypes**

The minor allele frequency (MAF) of the G allele in *rs10314* is 13.12% and of the G allele in rs885985 is 44.33% among those of EUR ancestry per LDpop, which references the 1000 Genomes Project (Alexander & Machiela, 2020). The minor alleles of *rs10314* is typically coinherited with rs885985 (D’ = .9864), but the two variants are not highly correlated (*R*^2^ = .1845 per LDlink; Machiela & Chanock, 2015), which suggests that associations observed with each are likely independent.

**Simoa Methods**

In the NCPTSD cohort, frozen plasma aliquots were shipped to the Quanterix Accelerator Lab (Quanterix Corporation, Billerica, MA) where the samples were processed per the manufacturer’s instructions. This included thawing and diluting samples X 4, and centrifugation for elimination of any particulates. Samples were run in 96 well plates with eight reference samples per plate. The four neurology markers were assayed on the Neurology 4-Plex E advantage kit in duplicate on the HD-X analyzer. The two inflammation markers (20% in duplicate) were obtained using the CorPlex Human Cytokine Panel 1 on the SP-X Imaging and Analysis System. In the TRACTS cohort, the plasma samples were analyzed (all in duplicate) on an in-house HD-1 analyzer (Quanterix, Billerica, MA). Aβ-42, Aβ-40, and GFAP and NFL were obtained on the Neurology 4-Plex A, and IL-6 and TNFα on the Cytokine 3-Plex A. Other Simoa methods are as described for the NCPTSD cohort. For both cohorts, samples that failed QC checks (e.g., unacceptable coefficient of variation [CV], poor calibration against reference samples) were rerun when possible. For both cohorts, samples were excluded if CV exceeded 25% or if the sample mean was greater than 3 SDs from the cohort mean. Samples with analytes measured above or below the functional upper or lower limits of quantification were set to the known respective upper and lower limits of quantification.

**RNA Methods**

Trimmomatic (Bolger et al., 2014) was applied to remove adapters and filter short and low-quality reads. The trimmed reads were then aligned to the hg38 human reference genome (Schneider et al., 2017) using STAR (Dobin et al., 2013). The qualities of the raw, trimmed, and aligned reads were checked using FastQC (https://www.bioinformatics.babraham.ac.uk/projects/fastqc/), RSeQC (Wang et al., 2012), and MultiQC (Ewels et al., 2016) separately. Samples were eliminated if there was evidence of less than 50% uniquely mapped reads. Transcriptome quantification was then performed with Kallisto (Bray et al., 2016), and the transcript abundance estimates were collapsed to the gene level using the tximport Bioconductor package. Gene expression values were log-transformed using the regularized log transformation (rLog) method implemented in DEseq2 (Love et al., 2014). Principal Components (PCs) were computed from the rLog-transformed expression values. Samples that were more than 6 standard deviations away from the mean on any of the first 10 PCs were considered outliers and excluded. For the remaining samples, self-report sex was confirmed by examining the expression of Y chromosome genes (Shi et al., 2016). Genotypes were called from the RNA-seq data using the GATK HaplotypeCaller (<https://gatk.broadinstitute.org/hc/en-us/articles/360037225632-HaplotypeCaller>) and compared with chip-based genotypes to confirm sample concordance. Estimates of cell type proportions in neural tissue were generated using BrainInABlender (Hagenauer et al., 2016).

**Mediation Model**

The mediation model (Figure 1) was conducted in Mplus 8.10 using the maximum likelihood estimator. NFL was regressed on age, sex, and cg21872764, which was, in turn, regressed on age, sex, 3 PCs, estimated white blood cells, and total number of types of traumatic life experiences. The model fit the data well. Trauma predicted cg21872764 (β = .120, p < .001) and cg21872764 predicted NFL levels (β = .077, p = .003) such that the indirect effect of trauma on NFL via cg21872764 was significant (β = .009, p = .015).

Table S1

*Pearson Correlations Among Three DNAm Probes Associated with PTSD Severity in Blood After FDR Correction (N = 1311)*

| Probe | cg00804504 | cg17411190 |
| --- | --- | --- |
| cg00804504 | -- |  |
| cg17411190 | -.230*** | -- |
| cg21872764 | -.158*** | .262*** |

****p* < .001

Please note, Table S2 is available as a separate excel file.

Table S3

*Associations Between CLDN5 SNPs, Trauma, PTSD, and CLDN5 DNAm Among European Participants (n = 873)*

|  | cg00804504 | | |  | cg17411190 | | |  | cg21872764 | | |
| --- | --- | --- | --- | --- | --- | --- | --- | --- | --- | --- | --- |
|  | B | β | *p* |  | B | β | *p* |  | B | β | *p* |
| Model 1 |  |  |  |  |  |  |  |  |  |  |  |
| rs10314 | .195 | .390 | < .00001 |  | -.075 | -.119 | .000027 |  | -.051 | -.074 | .003 |
| Trauma^a^ or PTSD Sev | -.075 | -.068 | .035 |  | .108 | .078 | .009 |  | .015 | .104 | .003 |
| Model 2 |  |  |  |  |  |  |  |  |  |  |  |
| rs885985 | .099 | .280 | < .00001. |  | -.107 | -.240 | < .00001 |  | -.067 | -.140 | .000026 |
| Trauma^a^ or PTSD Sev | -.078 | -.071 | .035 |  | .096 | .069 | .016 |  | .014 | .098 | .005 |

*Note*. Models covaried for age, sex, 3 ancestry substructure PCs, and estimated white blood cells.

^a^Trauma was evaluated in the model predicting cg21872764 only; PTSD severity was evaluated in the models predicting cg00804504 and cg17411190 because these are follow-up analyses to the models without the SNPs in which trauma exposure emerged as a predictor of cg21872764 and PTSD severity as a predictor of cg00804504 and cg17411190.

Table S4

*Associations between PTSD Diagnosis and all CLDN5 DNAm probes on the EPIC Chip in Postmortem Brain Tissue*

|  | dlPFC (*n* = 69) | | | vmPFC (*n* = 69) | | | Motor Cortex (*n* = 68) | | |
| --- | --- | --- | --- | --- | --- | --- | --- | --- | --- |
| CPG | B | β | *p* | B | β | *p* | B | β | *p* |
| cg17583256 | -0.143 | -0.136 | 0.335 | -0.162 | -0.136 | 0.265 | -0.023 | -0.025 | 0.865 |
| **cg17411190**^a^ | 0.114 | 0.129 | 0.381 | **0.305** | **0.369** | **0.005** | 0.196 | 0.238 | 0.148 |
| **cg09092054** | -0.173 | -0.194 | 0.220 | -0.164 | -0.145 | 0.359 | **0.404** | **0.472** | **0.001** |
| cg09446908 | 0.033 | 0.050 | 0.750 | -0.007 | -0.012 | 0.939 | -0.063 | -0.111 | 0.466 |
| cg17577122 | -0.107 | -0.084 | 0.590 | -0.226 | -0.182 | 0.252 | -0.338 | -0.290 | 0.058 |
| cg00189989 | -0.069 | -0.088 | 0.446 | -0.129 | -0.186 | 0.128 | 0.101 | 0.161 | 0.314 |
| cg16773741^a^ | -0.062 | -0.080 | 0.540 | -0.075 | -0.089 | 0.435 | 0.030 | 0.043 | 0.763 |
| cg00804504^a^ | -0.338 | -0.285 | 0.055 | -0.109 | -0.104 | 0.494 | 0.035 | 0.030 | 0.842 |
| cg00811132 | 0.047 | 0.050 | 0.747 | -0.048 | -0.052 | 0.740 | 0.151 | 0.133 | 0.371 |
| cg20486569 | 0.085 | 0.093 | 0.467 | 0.119 | 0.116 | 0.361 | 0.002 | 0.002 | 0.988 |
| **cg21872764**^a^ | 0.243 | 0.196 | 0.137 | **0.308** | **0.246** | **0.034** | 0.144 | 0.129 | 0.383 |
| cg04463638 | -0.044 | -0.062 | 0.572 | 0.094 | 0.137 | 0.181 | 0.139 | 0.230 | 0.061 |
| cg06340942 | 0.033 | 0.068 | 0.642 | 0.067 | 0.131 | 0.352 | 0.017 | 0.038 | 0.812 |
| cg11450827 | -0.044 | -0.064 | 0.645 | 0.043 | 0.063 | 0.586 | -0.059 | -0.093 | 0.556 |
| cg05498726 | 0.010 | 0.015 | 0.907 | 0.013 | 0.019 | 0.856 | -0.028 | -0.049 | 0.752 |
| cg05460329^a^ | -0.101 | -0.155 | 0.217 | 0.023 | 0.039 | 0.745 | 0.018 | 0.031 | 0.839 |
| cg13114849 | -0.097 | -0.179 | 0.215 | 0.019 | 0.032 | 0.807 | 0.151 | 0.289 | 0.063 |
| cg06315607 | 0.079 | 0.091 | 0.546 | -0.158 | -0.169 | 0.301 | -0.208 | -0.196 | 0.170 |
| cg14553765 | -0.135 | -0.198 | 0.112 | 0.060 | 0.082 | 0.495 | 0.065 | 0.102 | 0.486 |

*Note*. No probe was statistically significant after correction for multiple testing across all probes and brain regions. dlPFC = dorsolateral prefrontal cortex; vmPFC = ventromedial prefrontal cortex. ^a^Indicates the 3 corrected significant plus 2 nominally significant probes from our analyses in blood.

Table S5.

*Associations between CLDN5 DNAm and CLDN5 Expression in Postmortem Brain Tissue*

|  | dlPFC (*n* = 92) | | | vmPFC (*n* = 85) | | | Motor Cortex (*n* = 88) | | |
| --- | --- | --- | --- | --- | --- | --- | --- | --- | --- |
| CPG | B | β | *p* | B | β | *p* | B | β | *p* |
| cg17583256 | -0.014 | -0.015 | 0.801 | 0.023 | 0.029 | 0.756 | 0.075 | 0.081 | 0.279 |
| cg17411190^a^ | 0.034 | 0.030 | 0.596 | -0.036 | -0.029 | 0.734 | -0.001 | -0.001 | 0.994 |
| cg09092054 | -0.102 | -0.099 | 0.103 | -0.119 | -0.141 | 0.087 | -0.030 | -0.033 | 0.688 |
| **cg09446908** | -3.118E-04 | -2.180E-04 | 0.997 | **-0.343** | **-0.220** | **0.005** | -0.011 | -0.008 | 0.920 |
| **cg17577122** | 0.005 | 0.007 | 0.911 | **-0.118** | **-0.174** | **0.028** | -0.063 | -0.084 | 0.355 |
| cg00189989 | -0.098 | -0.076 | 0.221 | 0.134 | 0.100 | 0.236 | 0.048 | 0.038 | 0.620 |
| cg16773741^a^ | -0.092 | -0.073 | 0.288 | 0.064 | 0.058 | 0.516 | 0.116 | 0.090 | 0.282 |
| cg00804504^a^ | 0.053 | 0.068 | 0.244 | 0.109 | 0.140 | 0.117 | -0.018 | -0.025 | 0.759 |
| cg00811132 | 0.023 | 0.022 | 0.703 | 0.079 | 0.088 | 0.291 | -0.090 | -0.117 | 0.215 |
| cg20486569 | 0.007 | 0.007 | 0.906 | -0.007 | -0.007 | 0.933 | 0.028 | 0.026 | 0.761 |
| cg21872764^a^ | -0.007 | -0.008 | 0.888 | -0.073 | -0.092 | 0.292 | -0.030 | -0.038 | 0.637 |
| cg04463638 | 0.106 | 0.077 | 0.184 | 0.067 | 0.049 | 0.577 | 0.114 | 0.083 | 0.299 |
| cg06340942 | -0.098 | -0.054 | 0.369 | 0.155 | 0.096 | 0.253 | 0.117 | 0.064 | 0.428 |
| cg11450827 | -0.016 | -0.011 | 0.850 | 0.048 | 0.037 | 0.671 | 0.135 | 0.100 | 0.177 |
| cg05498726 | 0.004 | 0.002 | 0.969 | 0.121 | 0.090 | 0.324 | 0.174 | 0.123 | 0.132 |
| cg05460329^a^ | -0.069 | -0.047 | 0.457 | 0.090 | 0.061 | 0.486 | 0.038 | 0.027 | 0.744 |
| cg13114849 | -0.105 | -0.064 | 0.315 | 0.065 | 0.048 | 0.569 | -0.115 | -0.072 | 0.373 |
| cg06315607 | -0.026 | -0.025 | 0.691 | -0.098 | -0.106 | 0.196 | 0.057 | 0.069 | 0.410 |
| cg14553765 | -0.151 | -0.103 | 0.098 | 0.038 | 0.029 | 0.762 | 0.128 | 0.087 | 0.268 |

*Note*. dlPFC = dorsolateral prefrontal cortex; vmPFC = ventromedial prefrontal cortex. ^a^Indicates the 3 corrected significant plus 2 nominally significant probes from our analyses in blood. No associations were significant after multiple testing correction.

**References**

1000 Genomes Project Consortium, Auton, A., Brooks, L. D., Durbin, R. M., Garrison, E. P., Kang, H. M., Korbel, J. O., Marchini, J. L., McCarthy, S., McVean, G. A., & Abecasis, G. R. (2015). A global reference for human genetic variation. *Nature*, 526(7571), 68–74. <https://doi.org/10.1038/nature15393>

Alexander, T. A., & Machiela, M. J. (2020). LDpop: An interactive online tool to calculate and visualize geographic LD patterns. *BMC Bioinformatics*, 21(1), 14. <https://doi.org/10.1186/s12859-020-3340-1>

Aryee, M. J., Jaffe, A. E., Corrada-Bravo, H., Ladd-Acosta, C., Feinberg, A. P., Hansen, K. D., & Irizarry, R. A. (2014). Minfi: A flexible and comprehensive Bioconductor package for the analysis of Infinium DNA methylation microarrays. *Bioinformatics (Oxford, England)*, 30(10), 1363–1369. <https://doi.org/10.1093/bioinformatics/btu049>

Bolger, A. M., Lohse, M., & Usadel, B. (2014). Trimmomatic: A flexible trimmer for Illumina sequence data. *Bioinformatics (Oxford, England)*, 30(15), 2114–2120. <https://doi.org/10.1093/bioinformatics/btu170>

Bray, N. L., Pimentel, H., Melsted, P., & Pachter, L. (2016). Near-optimal probabilistic RNA-seq quantification. *Nature Biotechnology*, 34(5), 525–527. <https://doi.org/10.1038/nbt.3519>

Church, D. M., Schneider, V. A., Graves, T., Auger, K., Cunningham, F., Bouk, N., Chen, H. C., Agarwala, R., McLaren, W. M., Ritchie, G. R., Albracht, D., Kremitzki, M., Rock, S., Kotkiewicz, H., Kremitzki, C., Wollam, A., Trani, L., Fulton, L., Fulton, R., Matthews, L., … Hubbard, T. (2011). Modernizing reference genome assemblies. *PLoS Biology*, 9(7), e1001091. <https://doi.org/10.1371/journal.pbio.1001091>

Dobin, A., Davis, C. A., Schlesinger, F., Drenkow, J., Zaleski, C., Jha, S., Batut, P., Chaisson, M., & Gingeras, T. R. (2013). STAR: ultrafast universal RNA-seq aligner. *Bioinformatics (Oxford, England)*, 29(1), 15–21. <https://doi.org/10.1093/bioinformatics/bts635>

Ewels, P., Magnusson, M., Lundin, S., & Käller, M. (2016). MultiQC: Summarize analysis results for multiple tools and samples in a single report. *Bioinformatics (Oxford, England)*, 32(19), 3047–3048. <https://doi.org/10.1093/bioinformatics/btw354>

Hagenauer, M. H., Schulmann, A., Li, J. Z., Vawter, M. P., Walsh, D. M., Thompson, R. C., Turner, C. A., Bunney, W. E., Myers, R. M., Barchas, J. D., Schatzberg, A. F., Watson, S. J., & Akil, H. (2018). Inference of cell type content from human brain transcriptomic datasets illuminates the effects of age, manner of death, dissection, and psychiatric diagnosis. *PloS One*, 13(7), e0200003. <https://doi.org/10.1371/journal.pone.0200003>

Hastie, T., Tibshirani, R., Narasimhan, B., & Chu, G. (2023). *impute: Imputation for microarray data. R package version 1.74.1.*

Lam, M., Awasthi, S., Watson, H. J., Goldstein, J., Panagiotaropoulou, G., Trubetskoy, V., Karlsson, R., Frei, O., Fan, C. C., De Witte, W., Mota, N. R., Mullins, N., Brügger, K., Lee, S. H., Wray, N. R., Skarabis, N., Huang, H., Neale, B., Daly, M. J., Mattheisen, M., … Ripke, S. (2020). RICOPILI: Rapid Imputation for COnsortias PIpeLIne. *Bioinformatics (Oxford, England)*, 36(3), 930–933. <https://doi.org/10.1093/bioinformatics/btz633>

Leek, J. T., Johnson, W. E., Parker, H. S., Jaffe, A. E., & Storey, J. D. (2012). The sva package for removing batch effects and other unwanted variation in high-throughput experiments. *Bioinformatics (Oxford, England)*, 28(6), 882–883. <https://doi.org/10.1093/bioinformatics/bts034>

Love, M. I., Huber, W., & Anders, S. (2014). Moderated estimation of fold change and dispersion for RNA-seq data with DESeq2. *Genome Biology*, 15(12), 550. <https://doi.org/10.1186/s13059-014-0550-8>

Machiela, M. J., & Chanock, S. J. (2015). LDlink: A web-based application for exploring population-specific haplotype structure and linking correlated alleles of possible functional variants. *Bioinformatics (Oxford, England)*, 31(21), 3555–3557. <https://doi.org/10.1093/bioinformatics/btv402>

Ratanatharathorn, A., Boks, M. P., Maihofer, A. X., Aiello, A. E., Amstadter, A. B., Ashley-Koch, A. E., Baker, D. G., Beckham, J. C., Bromet, E., Dennis, M., Garrett, M. E., Geuze, E., Guffanti, G., Hauser, M. A., Kilaru, V., Kimbrel, N. A., Koenen, K. C., Kuan, P. F., Logue, M. W., Luft, B. J., … Smith, A. K. (2017). Epigenome-wide association of PTSD from heterogeneous cohorts with a common multi-site analysis pipeline. *American Journal of Medical Genetics Part B: Neuropsychiatric Genetics,* 174(6), 619–630. <https://doi.org/10.1002/ajmg.b.32568>

Schneider, V. A., Graves-Lindsay, T., Howe, K., Bouk, N., Chen, H. C., Kitts, P. A., Murphy, T. D., Pruitt, K. D., Thibaud-Nissen, F., Albracht, D., Fulton, R. S., Kremitzki, M., Magrini, V., Markovic, C., McGrath, S., Steinberg, K. M., Auger, K., Chow, W., Collins, J., Harden, G., … Church, D. M. (2017). Evaluation of GRCh38 and de novo haploid genome assemblies demonstrates the enduring quality of the reference assembly. *Genome Research*, 27(5), 849–864. <https://doi.org/10.1101/gr.213611.116>

Shi, L., Zhang, Z., & Su, B. (2016). Sex biased gene expression profiling of human brains at major developmental stages. *Scientific Reports*, 6, 21181. <https://doi.org/10.1038/srep21181>

Wang, L., Wang, S., & Li, W. (2012). RSeQC: quality control of RNA-seq experiments. *Bioinformatics (Oxford, England)*, 28(16), 2184–2185. <https://doi.org/10.1093/bioinformatics/bts356>
